# The Burden of Cardiovascular Events According to Cardiovascular Risk Profile in Adults from High-, Middle- and Low-Income Countries

**DOI:** 10.1101/2024.03.11.24304132

**Authors:** Darryl P. Leong, Rita Yusuf, Romaina Iqbal, Álvaro Avezum, Afzalhussein Yusufali, Annika Rosengren, Jephat Chifamba, Fernando Lanas, Maria Luz Diaz, J. Jaime Miranda, Kairat Davletov, Erkin Mirrakhimov, Karen Yeates, Rasha Khatib, Khalid F. Alhabib, Sadi Gulec, Maria José Paucar, Jose Patricio Lopez-Lopez, Salim Yusuf

**Affiliations:** Population Health Research Institute, McMaster University and Hamilton Health Sciences, Hamilton, Canada; School of Environment and Life Sciences, Independent University, Bangladesh, Dhaka, Bangladesh; Department of Community Health Sciences, Aga Khan University, Karachi, Pakistan; International Research Center, Hospital Alemão Oswaldo Cruz, São Paolo, Brazil; Tamami Foundation, Matemwe, Zanzibar, Tanzania; Sahlgrenska Academy, University of Gothenburg and Sahlgrenska University Hospital, VGR Region, Sweden; Department of Biomedical Sciences, University of Zimbabwe, Harare, Zimbabwe; Universidad de la Frontera, Temuco, Chile; Estudios Clinicos Latinoamerica, Instituto Cardiovascular de Rosario, Rosario, Argentina; Centro de Excelencia en Enfermedades Crónicas, Universidad Peruana Cayetano Heredia, Lima, Perú; Asfendiarov Kazakh National Medical University, Almaty, Kazakhstan; Kyrgyz State Medical Academy and National Center of Cardiology and Internal Disease, Bishkek, Kyrgyzstan; Department of Medicine, Queen’s University, Kingston, Canada; Advocate Aurora Research Institute, Advocate Aurora Health, Milwaukee, USA; Department of Cardiac Sciences, King Fahad Cardiac Center, King Saud University, Riyadh, Saudi Arabia; Cardiology Department, Ankara University School of Medicine, Ankara, Türkiye; Facultad de Ciencias de la Salud Eugenio Espejo, Universidad UTE, Quito, Ecuador; Masira Research Institute, Universidad de Santander, Bucaramanga, Colombia

## Abstract

**Background:** Current strategies to prevent adverse cardiovascular outcomes focus on secondary prevention and primary prevention in high-risk groups. The proportion of events occurring in lower-risk groups globally is unknown.

**Methods:** We prospectively documented fatal or non-fatal myocardial infarction, stroke, heart failure, or any other fatal cardiovascular event stratified by history of cardiovascular disease (CVD), and by the INTERHEART and the Framingham risk score in those without prior CVD, in 189,097 adults from 26 high-, middle- and low-income countries.

**Results:** Participants’ mean±SD age was 51±10 years and 59% were women. We observed 14,829 outcome events affecting 8% of the cohort during a median 12.4 years’ follow-up. Overall, 44% of outcome events occurred in CVD-naïve participants at low or intermediate INTERHEART risk and 56% occurred in in CVD-naïve participants at non-high Framingham risk. The proportion of adverse cardiovascular outcomes occurring in these lower risk groups was inversely related to country income level and was higher in women (55%) than in men (35%).

**Conclusions:** To achieve a substantial population-level reduction in CVD, preventive strategies for CVD are essential in those considered low- or intermediate-risk.

## INTRODUCTION

Currently, strategies for the prevention of adverse cardiovascular outcomes focus on secondary prevention(1) and primary prevention in high-risk groups(2). However, in European populations, most cardiovascular events occur in those with no history of cardiovascular disease and without established cardiovascular risk factors because most of the population falls into this category(3). Therefore, it has been postulated that population-wide approaches to preventing cardiovascular events would lead to greater reductions in these outcomes than the current approach of targeting only those at high-risk for or with established cardiovascular disease(4,5).

Most cardiovascular events globally occur in low- and middle-income countries(6). However, there is a paucity of data from these countries on the occurrence of cardiovascular events in those with established cardiovascular disease or at high cardiovascular risk versus those at low cardiovascular risk. Therefore, in this analysis of the Prospective Urban Rural Epidemiology (PURE) study, our aim was to describe the proportion of adverse cardiovascular events stratified by baseline cardiovascular risk in adults from low-, middle- and high-income countries.

## METHODS

The PURE study is an ongoing, international, prospective, population-based cohort study whose design has previously been described(7,8). The study is conducted in high- (HIC), middle- (MIC) and low-(LIC) income countries based on gross national income per capita according to the World Bank classification in 2006 when most countries were recruiting. This analysis includes 189,097 participants from 26 countries who had at least one follow-up visit. The HIC were Canada (n=10,462), Saudi Arabia (n=2046), Sweden (n=4153) and United Arab Emirates (n=1499). The MIC were Argentina (n=7534), Brazil (n=6081), Chile (n=3588), China (n=47,732), Colombia (n=7545), Ecuador (n=2027), Iran (n=6012), Kazakhstan (n=2176), Kyrgyzstan (n=1125), Malaysia (n=15,789), Palestine (n=1668), Philippines (n=5006), Poland (n=14,790), Russia (n=13572), Türkiye (n=4059) and South Africa (n=4533). The LIC were Bangladesh (n=2936), India (n=28,932), Pakistan (n=2397), Peru (n=2327), Tanzania (n=2050) and Zimbabwe (n=1258).

In each participating country, individuals were recruited from households in urban (n=102,187) and rural (n=86,910) communities. Households were selected to be broadly representative of the socio-demographic composition of communities. Although not designed to be nationally representative, the socio-demographic characteristics and death rates of men and women were similar to their respective national populations^8^.

The PURE study received approval from the ethics committees at each centre, and all participants provided written informed consent.

### Data collection

At baseline, standardized questionnaires were used to collect information on participant characteristics and medical history. Participants were considered to have a past history of cardiovascular disease if they reported being diagnosed with angina, myocardial infarction, coronary artery disease, stroke or heart failure. Diet quality was evaluated by the Alternative Health Eating Index, whereby a higher score is associated with a lower risk of future (VDC9). Physical activity was assessed using the International Physical Activity Questionnaire(10). Depression was evaluated by asking whether the participant felt sad, blue or depressed for at least two consecutive weeks during the past twelve months. Stress was assessed by asking how often the participant felt stressed at home and (if still working) at work during the past twelve months.

Physical measurements were performed using standardized equipment and procedures. Blood pressure was measured with the participant sitting for 5 minutes using an automated sphygmomanometer (OMRON Healthcare Inc, Toronto). Weight was measured using a digital scale (Tanita Corporation) with the participant lightly clothed with no shoes. Height was measured using a tape measure with the participant standing without shoes. Waist and hip circumferences were measured unclothed using a tape measure. The waist circumference was considered the smallest circumference between the costal margin and the iliac crest. Blood was collected at baseline and total and high-density lipoprotein (HDL-C) cholesterol were measured.

In participants without a history of CVD, we calculated the non-laboratory INTERHEART cardiovascular risk score, which is a validated measure of risk for future myocardial infarction based on age, smoking, second hand smoke exposure, self-reported diabetes and hypertension, family history of myocardial infarction, waist-hip ratio, dietary habits, physical activity levels and self-reported stress and depression(11). The INTERHEART score is measured on a scale of 0 to 48 with higher scores indicating higher risk. In addition, the Framingham cardiovascular risk score was calculated from participant age, systolic blood pressure, blood pressure treatment, smoking and diabetes, total cholesterol and HDL-C. Framingham score was considered high if >14 in men and >17 in women, as these thresholds are associated with >20% 10-year risk of a CVD event(12).

### Outcomes

Participants were contacted at least every three years to ascertain their vital status and the occurrence of specific non-fatal events. Where possible, detailed information on cause of death was obtained from verbal autopsies or medical records, and on non-fatal events from hospital or physician reports and standardized questionnaires. The primary outcome of this analysis was a new CVD event, defined as fatal or non-fatal myocardial infarction, stroke, heart failure, or any other fatal event determined to be of cardiovascular cause. These events were adjudicated centrally in each country by trained physicians using standardized definitions (Supplementary Material).

### Statistical Analysis

Participants were stratified into those with a history of cardiovascular disease, and in those without a history of cardiovascular disease, by tertile of INTERHEART risk score. As a sensitivity analysis, we repeated analyses stratifying those without a history of cardiovascular disease by according to their Framingham risk score.

## RESULTS

Participant baseline characteristics are displayed in Table 1. Mean±standard deviation age was 51±10 years. Women, who comprised 59% of the cohort, had lower INTERHEART risk scores than men and at baseline, 6892 (6.2%) women had a history of CVD, as compared with 5254 (6.8%) men (p<0.0001). There was a positive association between INTERHEART risk score and each of country income level, education and current alcohol use. In HIC, MIC and LIC respectively, baseline CVD was present in 969 (5.3%); 9949 (7.6%); and 1228 (3.1%) (p<0.0001). Among those with primary, secondary, and beyond secondary school education, 4523 (6.1%), 3465 (5.3%) and 2374 (6.6%) had a history of CVD respectively (p<0.0001). Among alcohol never drinkers, current drinkers, and former drinkers, 6890 (5.7%); 3542 (6.6%); and 1239 (13.6%) had a history of CVD respectively (p<0.0001).

**Table 1.** Participant baseline characteristics. AHEI = alternate healthy eating index; IHRS = non-laboratory INTERHEART risk score. P-values across the four mutually exclusive groups were all <0.001, indicating values for each characteristic differed among these groups.

| Characteristic | No prior CVD |  |  | Baseline CVD<br>N=12,146 |
| --- | --- | --- | --- | --- |
|  | Low IHRS<br>N=56,967 | Intermediate IHRS<br>N=68,053 | High IHRS<br>N=51,927 |  |
| Age, years | 47±9 | 51±10 | 54±9 | 57±9 |
| Female | 41,974 (74) | 40,659 (60) | 22,505 (43) | 6892 (57) |
| Country income level |  |  |  |  |
| High | 2573 (4) | 6329 (9) | 8289 (16) | 969 (8) |
| Middle | 35,809 (63) | 48,010 (71) | 37,269 (72) | 9949 (82) |
| Low | 18,589 (33) | 13,714 (20) | 6369 (12) | 1228 (10) |
| Education |  |  |  |  |
| Primary | 23,372 (44) | 26,414 (42) | 19,503 (40) | 4523 (44) |
| Secondary | 24,667 (39) | 24,027 (38) | 17,752 (36) | 3465 (33) |
| Post-secondary | 9018 (17) | 13,077 (20) | 11,650 (24) | 2374 (23) |
| Tobacco use |  |  |  |  |
| Never | 47,464 (85) | 46,448 (69) | 23,696 (46) | 7136 (60) |
| Former | 2989 (5) | 8386 (12) | 10,938 (21) | 2668 (23) |
| Current | 5386 (10) | 12,853 (19) | 17,190 (33) | 1992 (17) |
| Alcohol use |  |  |  |  |
| Never | 43,253 (79) | 44,138 (66) | 26,310 (52) | 6890 (59) |
| Former | 1386 (3) | 2994 (5) | 3518 (7) | 1239 (11) |
| Current | 9748 (18) | 19,546 (29) | 21,126 (41) | 35421 (30) |
| Diabetes (self-reported) | 84 (<1) | 2429 (4) | 10,204 (20) | 2591 (21) |
| Glucose, mmol/L | 4.8±0.8 | 5.1±1.4 | 6.0±2.7 | 5.7±2.4 |
| Hypertension (self-reported) | 2957 (5) | 15,868 (19) | 19,608 (38) | 7122 (59) |
| Systolic blood pressure, mmHg | 119±14 | 132±21 | 142±22 | 139±24 |
| Body-mass index, kg/m<sup>2</sup> | 24.3±5.1 | 25.9±5.3 | 27.8±5.7 | 27.6±5.8 |
| Waist-hip ratio | 0.83±0.07 | 0.87±0.08 | 0.92±0.08 | 0.90±0.09 |
| AHEI score | 35.2±7.7 | 35.0±8.2 | 33.8±8.7 | 35.5±8.5 |
| Physical activity |  |  |  |  |
| Low | 5953 (11) | 11,575 (18) | 11,335 (23) | 2131 (18) |
| Medium | 21,092 (41) | 23,488 (37) | 16,442 (34) | 4690 (41) |
| High | 24,748 (48) | 28,016 (45) | 20,841 (43) | 4732 (41) |
| Depressed | 5315 (10) | 12,176 (18) | 14,585 (28) | 3379 (28) |
| Home stress |  |  |  |  |
| Never | 20,337 (50) | 24,488 (44) | 17,038 (38) | 3596 (42) |
| Some periods | 18,360 (46) | 23,807 (43) | 17,678 (39) | 3596 (38) |
| Several periods | 1321 (3) | 5495 (10) | 7350 (16) | 1173 (13) |
| Permanent | 471 (1) | 2025 (3) | 3125 (7) | 636 (7) |
| Work stress |  |  |  |  |
| Never | 13,147 (49) | 14,897 (41) | 9448 (33) | 1812 (40) |
| Some periods | 12,473 (46) | 15,482 (43) | 10,549 (37) | 1730 (36) |
| Several periods | 984 (4) | 4384 (12) | 5956 (21) | 624 (14) |
| Permanent | 393 (1) | 1623 (4) | 2680 (9) | 341 (8) |

Among never smokers, current smokers, and former smokers, 7136 (5.7%), 1992 (5.3%); and 2668 (10.7%) had a history of CVD respectively (p<0.0001). Of individuals with versus without diabetes, there were 2591 (16.9%) versus 9525 (5.5%) with known CVD (p<0.0001). Of individuals with versus without hypertension, there were 7122 (16.7%) versus 4942 (3.4%) with known CVD (p<0.0001). Respective body-mas indices and waist-hip ratios in those with versus without baseline CVD were 27.6±5.8kg/m^2^ versus 26.0±5.5kg/m^2^ (p<0.0001); and 0.90±0.09 versus 0.88±0.09 (p<0.0001). Those with a history of CVD had healthier diets but were less physically active and had higher rates of depression and stress than those who did not have known CVD.

### Outcomes

During a median 12.4 years’ follow-up, 14,829 (8%) of participants experienced a major adverse cardiovascular event (myocardial infarction, stroke, heart failure, or other fatal CVD). The proportion of incident CVD events occurring in four mutually exclusive groups – those with baseline CVD, and those without baseline CVD stratified by tertile of INTERHEART risk score – is shown in Figure 1. Overall, nearly half of incident CVD occurred in individuals with no prior history of CVD and with a low or intermediate INTERHEART risk score. There was a progressive increase in the proportion of CVD occurring in low or intermediate risk groups from HIC to MIC to LIC (Figure 2). In HIC, approximately one-fifth of CVD events occurred in those with no known CVD and with a low or intermediate INTERHEART risk score. This proportion increased to approximately two-fifths in MIC, and to nearly three-fifths in LIC.

**Figure 1.**
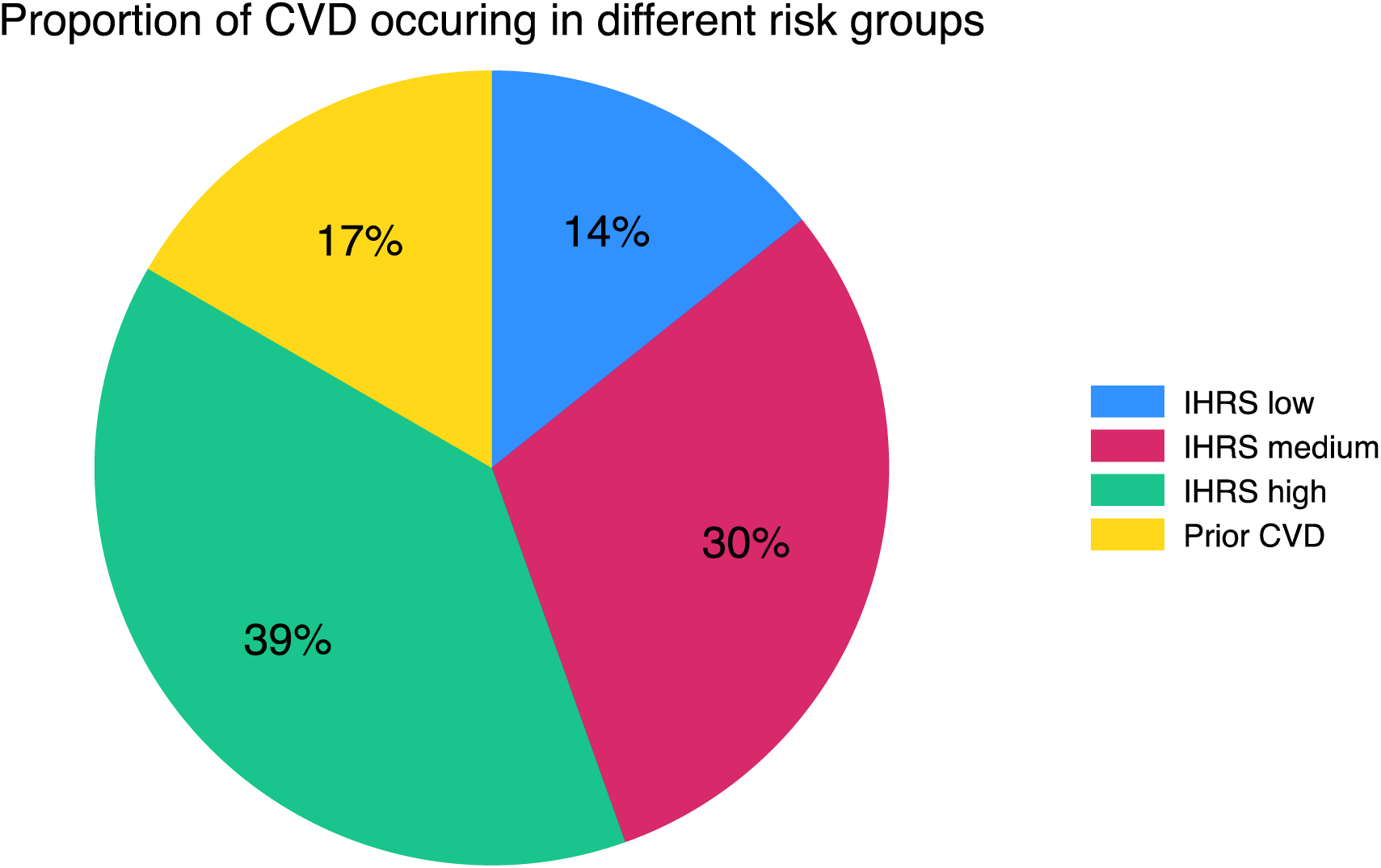
Proportion of incident cardiovascular disease (CVD) events occurring in different cardiovascular risk groups. Incident CVD includes myocardial infarction, stroke, heart failure or other fatal cardiovascular event. The four mutually exclusive cardiovascular risk groups were: past history of non-fatal CVD and in those with no past history of CVD, according to thirds of INTERHEART risk score (IHRS).

**Figure 2.**
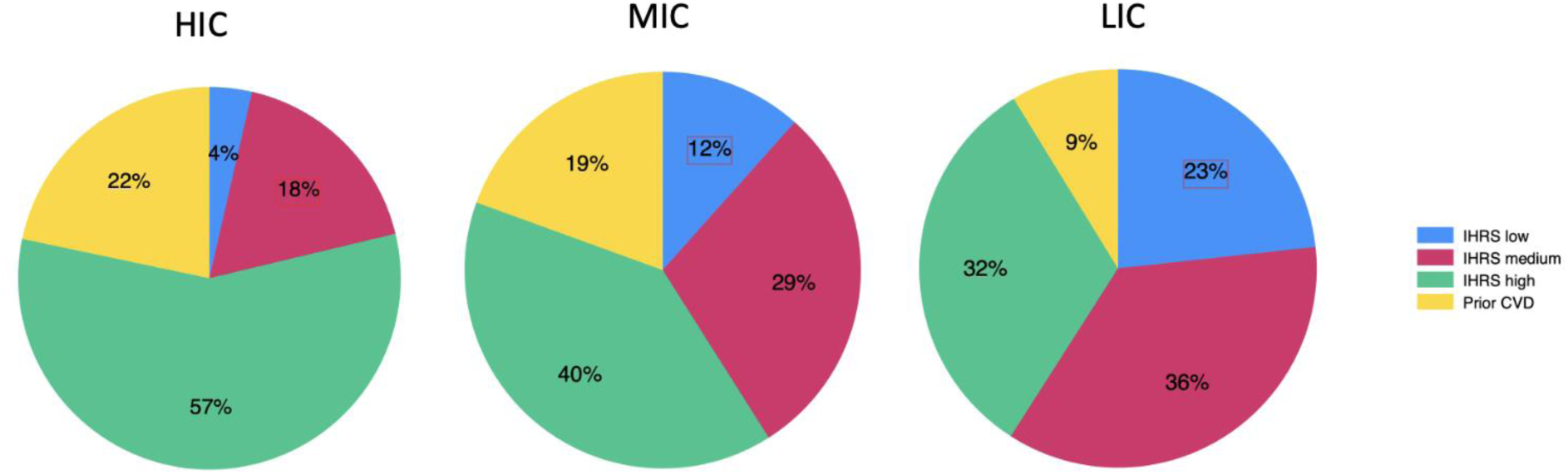
Proportion of incident cardiovascular disease (CVD) events occurring in different cardiovascular risk groups stratified by high (HIC)-, middle (MIC)-, and low (LIC)-income country level. Incident CVD includes myocardial infarction, stroke, heart failure or other fatal cardiovascular event. The four mutually exclusive cardiovascular risk groups were: past history of non-fatal CVD and in those with no past history of CVD, according to thirds of INTERHEART risk score (IHRS).

Overall, 56% of CVD events occurred in those whose Framingham risk score was low or intermediate, 28% in those with a high Framingham risk score and 17% in those with a past history of CVD (Figure 3). In HIC, 51% of CVD events occurred in those whose Framingham risk score was low or intermediate, 27% in those with a high Framingham risk score and 22% in those with a past history of CVD. Respective percentages in MIC and LIC were 54%, 27% and 19%; and 62%, 29% and 9%.

**Figure 3.**
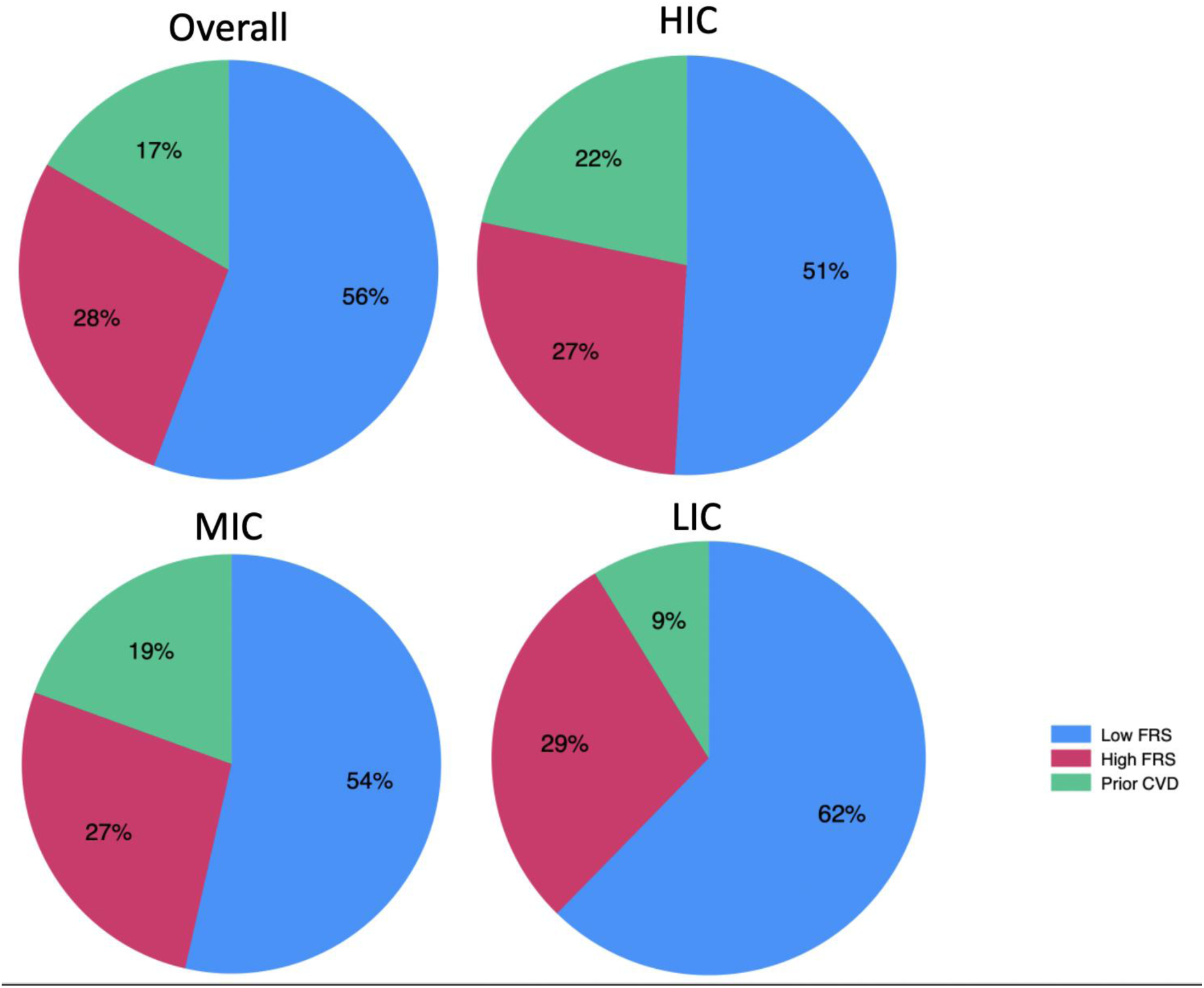
Proportion of incident cardiovascular disease (CVD) events occurring in different cardiovascular risk groups, both overall and stratified by high (HIC)-, middle (MIC)-, and low (LIC)-income country level. Incident CVD includes myocardial infarction, stroke, heart failure or other fatal cardiovascular event. The three mutually exclusive cardiovascular risk groups were: past history of non-fatal CVD and in those with no past history of CVD, high Framingham risk score (>14 in men or >17 in women) versus non-high.

We observed differences between men and women in the proportions of CVD events occurring in lower INTERHEART risk groups (Figure 4). In women, most (55%) CVD developed in those with no known CVD who were classified at low or intermediate risk, as compared with men, among whom 35% of CVD occurred among low or intermediate risk individuals. Among women, the proportion of CVD events occurring in those with no known CVD who were classified at low risk was inversely related to country income level, with 6%, 17% and 30% of events occurring in low-risk individuals in HIC, MIC and LIC respectively. A similar pattern was seen in men, with 2%, 6% and 18% of CVD events occurring in those at low risk with no known CVD in HIC, MIC and LIC respectively. Similar patterns were seen when implementing the Framingham risk score for the INTERHEART score (data not shown).

**Figure 4.**
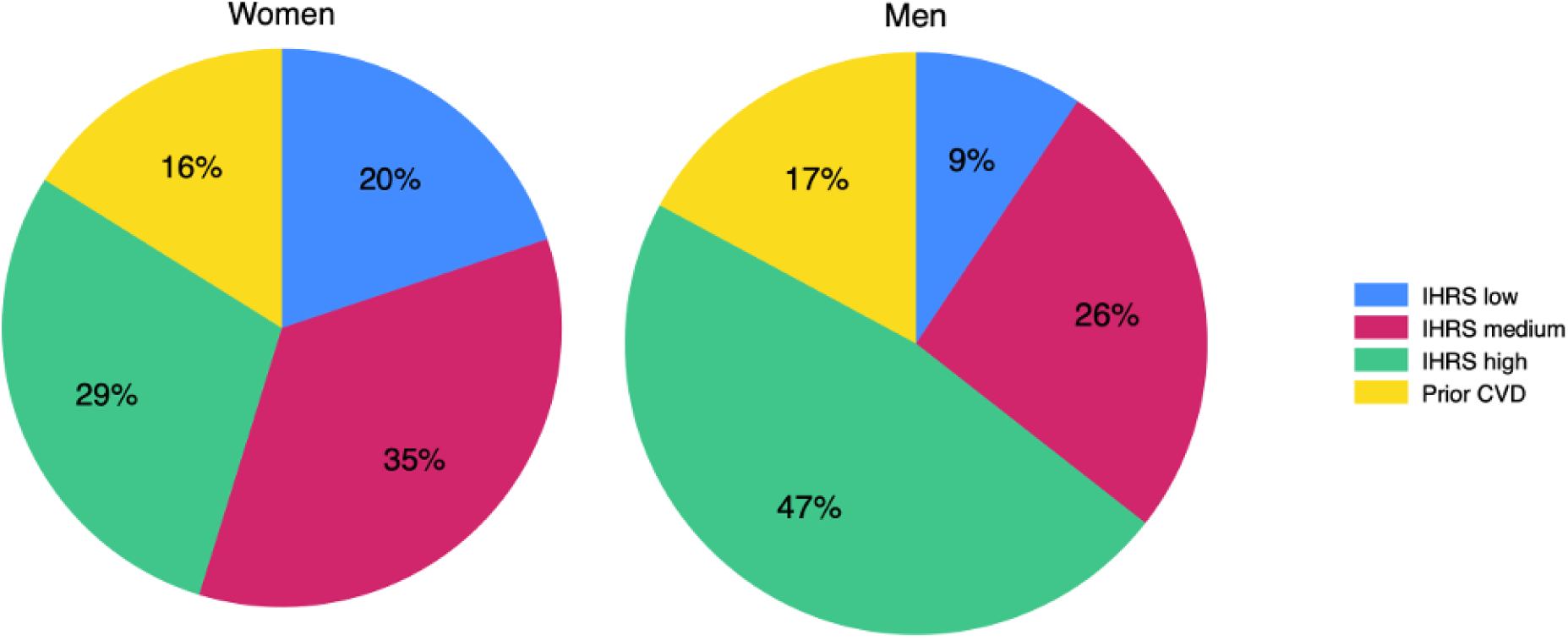
Proportion of incident cardiovascular disease (CVD) events occurring in different cardiovascular INTERHEART risk groups (IHRS) stratified by sex.

We examined the proportion of adverse cardiovascular events occurring in the presence of the major individual CVD risk factors. Of all incident CVD events, 23% developed in those with diabetes (self-reported, on blood glucose-lowering medications or with fasting plasma glucose level ≥7mmol/L) and 26% developed in current smokers.

In total, 39% of CVD events occurred in those with self-reported hypertension. However, there were 37,771 participants who reported not having hypertension but who had baseline systolic blood pressure ≥140mmHg and/or diastolic blood pressure ≥90mmHg. This represents 26% of individuals who reported not having hypertension. When these individuals are included in those with baseline hypertension, we observed that 65% of CVD occurred in this wider group. In both HIC and MIC, 69% of CVD events occurred in participants with self-reported hypertension or baseline systolic blood pressure ≥140mmHg and/or diastolic blood pressure ≥90mmHg, while this proportion was 54% in LIC.

Overall, 59% of CVD events occurred in those aged >55 years. The respective proportions of CVD events occurring in those >55 years in HIC, MIC and LIC were 67%, 61% and 53%.

## DISCUSSION

The main findings from this analysis of adults from 26 HIC, MIC and LIC are that: 1) overall, approximately half of severe CVD events develop in individuals with no history of CVD and who are not considered at high CVD risk according to validated risk scores; 2) the proportion of CVD events occurring in these low- and intermediate-risk individuals doubles from HIC to MIC and then again from MIC to LIC; 3) the burden of CVD events developing in lower-risk individuals is greater among women than men; and 4) among the traditional CVD risk factors, only hypertension (including unrecognized hypertension) identified a group of individuals that accounted for the majority of major CVD events during follow-up.

### Cardiovascular Risk and the Burden of Incident Severe CVD Events

In a cohort of Italian adults without CVD aged 35-69 years at enrolment, the overwhelming majority (89%) of incident cases of stroke occurred in those with a high burden of cardiovascular risk factors(13). In contrast, other European and American population-based cohort data suggest that most major CVD events occur in individuals not considered at high cardiovascular risk based on traditional CVD risk factors(14). Our findings are consistent with the latter data as we found that at approximately half of cases of incident severe CVD occurred in those with a low or intermediate burden of cardiovascular risk factors. There are several reasons that might account for these divergent findings. In the Italian study, baseline data collection occurred between 1983 and 1997 versus post-2001 in the PURE study. There have been some (albeit modest) improvements in the uptake of cardiovascular medications during the past 4 decades. Consistent with this, 11% of the Italian cohort were taking antihypertensive medication as compared with 21% in HIC in the PURE cohort.

Improvements in the treatment of cardiovascular risk factors may reduce the risk of major CVD in high-risk individuals. Therefore, we may be witnessing an epidemiologic transition in that as individuals at high-risk increasingly receive preventive treatments, the relative burden of adverse cardiovascular events may shift towards low-risk individuals who do not receive these treatments.

Contemporary recommendations indicate that individuals at low cardiovascular risk should only be counselled on healthy lifestyle, while the addition of pharmacotherapy to lifestyle advice should be reserved for those at higher cardiovascular risk(15,16). These recommendations are axiomatic in that they are widely accepted and advocated. However, Wald and Law argued that as 96% of deaths from ischemic heart disease and stroke occur in adults aged ≥55 years, and the risks of preventive medications like statins, antihypertensive medications and aspirin are small, all adults ≥55 years should receive a polypill containing these medications(5). We found that nearly two-thirds of fatal and non-fatal CVD events occurred in those aged ≥55 years. These data may be helpful in modelling the effects of Wald and Law’s hypothesis.

The World Heart Federation declared a target of 30 by 30 – i.e. a 30% decrease in cardiovascular disease and death by 2030. CVD is the commonest cause of death globally(8,17). While cardiovascular death is to a large extent preventable, to achieve a 30% reduction in cardiovascular disease and death, large reductions in deaths from CVD would be required. A major modelling exercise published in 2016 found that it is possible to achieve an aligned goal of 25 by 25 if there is a population-wide 25% decrease in the prevalence of systolic blood pressure >140mmHg, a 30% reduction in smoking, and no increases in fasting plasma glucose and BMI(18). While some of these milestones may have been achieved in select settings (e.g. decreasing smoking rates in North America)(19), others (e.g. blood pressure targets) have not(20,21). Unless a reduction in CVD events occurs in those at low- or intermediate-risk, it is unlikely that 30 by 30 will be realized.

### The Disproportionate Burden of CVD Occurring in Women at Lower CVD Risk

Among women with no known CVD in the Chicago Heart Association Detection Project in Industry Study, only 15% of cardiovascular deaths occurred in those at lower CVD risk(22). Our findings in HIC are consistent with this. Our study extends on these results by demonstrating that in women from MIC and LIC, low-CVD risk groups accounted for 3-fold and 5-fold more adverse cardiovascular outcomes than in HIC.

We have previously shown that women in the PURE study have a lower CVD risk factor burden than men and among those without established CVD, more primary prevention medication use(23). The present analysis extends on this to demonstrate that despite these favourable characteristics, a larger burden of adverse cardiovascular outcomes was observed in women at lower CVD risk, especially in MIC and LIC. These findings suggest that other non-traditional and/or sex-specific risk factors might account for the excess of cardiovascular events in women considered low-risk.

### Hypertension

We found that most major CVD developed in individuals with established hypertension or who had high blood pressure but were unaware of having hypertension. This observation is consistent with analyses from the Global Burden of Disease suggesting that among cardiovascular risk factors, hypertension accounts for the largest number of deaths(24). The contribution of hypertension to CVD incidence is related to both inadequate blood pressure treatment in many individuals diagnosed with hypertension and to under-diagnosis of hypertension. Given the large burden of hypertension, innovative strategies, such as the deployment of physicians or non-physician healthcare workers specifically to address hypertension may be helpful in achieving better blood pressure in communities(25,26). Fixed-dose combination “polypills” have also been proven effective in decreasing blood pressure in adults with intermediate levels of cardiovascular risk(27). Improving access to CVD medicines should be a key part of the strategy to lower CVD burden globally(28).

### Strengths and Limitations

One limitation of this analysis is that cardiovascular risk was only evaluated at baseline; we did not account for change in cardiovascular risk during follow-up. Also, the prognostic value of the Framingham risk score has not been validated in all the countries included in our analysis. However, it has been shown to have good discriminatory value in some of the included countries, such as Malaysia(29). The INTERHEART score was developed for the prediction of myocardial infarction, not stroke or heart failure, which were included in the cardiovascular composite outcome in this study. However, the Framingham score demonstrates good discriminatory ability for coronary heart disease, cerebrovascular disease and heart failure(30).

The main strength of this analysis is that to our knowledge, there are no single studies that have recorded cardiovascular risk factors and CVD outcomes in a standardized manner in a cohort of this size in countries of all income levels. The ascertainment of outcome events was performed using a rigorous process.

### Conclusions

Most major CVD events occur in adults with no history of CVD and who are not considered at high cardiovascular risk. This is most marked in low-income countries. To achieve a substantial population-level reduction in CVD, preventive strategies for CVD are essential in those considered low- or intermediate-risk.

## Data Availability

Individual-level data will not be made publicly available because further analyses are planned using the data and because consent has not been obtained from study participants to release these data to researchers who are not study investigators. On reasonable request, aggregate data will be made available. Study case report forms, the laboratory procedures manual and statistical code will also be shared on request.

## REFERENCES

1. Fihn SD, Gardin JM, Abrams J et al. 2012 ACCF/AHA/ACP/AATS/PCNA/SCAI/STS Guideline for the diagnosis and management of patients with stable ischemic heart disease: a report of the American College of Cardiology Foundation/American Heart Association Task Force on Practice Guidelines, and the American College of Physicians, American Association for Thoracic Surgery, Preventive Cardiovascular Nurses Association, Society for Cardiovascular Angiography and Interventions, and Society of Thoracic Surgeons. J Am Coll Cardiol 2012;60:e44–e164.

2. Arnett DK, Blumenthal RS, Albert MA et al. 2019 ACC/AHA Guideline on the Primary Prevention of Cardiovascular Disease: Executive Summary: A Report of the American College of Cardiology/American Heart Association Task Force on Clinical Practice Guidelines. J Am Coll Cardiol 2019;74:1376–1414.

3. Cooney MT, Dudina A, Whincup P et al. Re-evaluating the Rose approach: comparative benefits of the population and high-risk preventive strategies. Eur J Cardiovasc Prev Rehabil 2009;16:541–9.

4. Rose G. Sick individuals and sick populations. Int J Epidemiol 2001;30:427–32; discussion 433-4.

5. Wald NJ, Law MR. A strategy to reduce cardiovascular disease by more than 80%. BMJ 2003;326:1419.

6. Roth GA, Mensah GA, Johnson CO et al. Global Burden of Cardiovascular Diseases and Risk Factors, 1990-2019: Update From the GBD 2019 Study. J Am Coll Cardiol 2020;76:2982–3021.

7. Corsi DJ, Subramanian SV, Chow CK et al. Prospective Urban Rural Epidemiology (PURE) study: Baseline characteristics of the household sample and comparative analyses with national data in 17 countries. Am Heart J 2013;166:636–646 e4.

8. Dagenais GR, Leong DP, Rangarajan S et al. Variations in common diseases, hospital admissions, and deaths in middle-aged adults in 21 countries from five continents (PURE): a prospective cohort study. Lancet 2020;395:785–794.

9. Dehghan M, Mente A, Teo KK et al. Relationship between healthy diet and risk of cardiovascular disease among patients on drug therapies for secondary prevention: a prospective cohort study of 31 546 high-risk individuals from 40 countries. Circulation 2012;126:2705–12.

10. Lear SA, Hu W, Rangarajan S et al. The effect of physical activity on mortality and cardiovascular disease in 130 000 people from 17 high-income, middle-income, and low-income countries: the PURE study. Lancet 2017;390:2643–2654.

11. McGorrian C, Yusuf S, Islam S et al. Estimating modifiable coronary heart disease risk in multiple regions of the world: the INTERHEART Modifiable Risk Score. Eur Heart J 2011;32:581–9.

12. Anderson TJ, Gregoire J, Pearson GJ et al. 2016 Canadian Cardiovascular Society Guidelines for the Management of Dyslipidemia for the Prevention of Cardiovascular Disease in the Adult. Can J Cardiol 2016;32:1263–1282.

13. Giampaoli S, Palmieri L, Panico S et al. Favorable cardiovascular risk profile (low risk) and 10-year stroke incidence in women and men: findings from 12 Italian population samples. Am J Epidemiol 2006;163:893–902.

14. Polonsky TS, Greenland P. CVD screening in low-risk, asymptomatic adults: clinical trials needed. Nat Rev Cardiol 2012;9:599–604.

15. Wong ND, Budoff MJ, Ferdinand K et al. Atherosclerotic cardiovascular disease risk assessment: An American Society for Preventive Cardiology clinical practice statement. Am J Prev Cardiol 2022;10:100335.

16. Arnett DK, Blumenthal RS, Albert MA et al. 2019 ACC/AHA Guideline on the Primary Prevention of Cardiovascular Disease: A Report of the American College of Cardiology/American Heart Association Task Force on Clinical Practice Guidelines. Circulation 2019;140:e596–e646.

17. Collaborators GBDCoD. Global, regional, and national age-sex-specific mortality for 282 causes of death in 195 countries and territories, 1980-2017: a systematic analysis for the Global Burden of Disease Study 2017. Lancet 2018;392:1736–1788.

18. Sacco RL, Roth GA, Reddy KS et al. The Heart of 25 by 25: Achieving the Goal of Reducing Global and Regional Premature Deaths From Cardiovascular Diseases and Stroke: A Modeling Study From the American Heart Association and World Heart Federation. Circulation 2016;133:e674–90.

19. Dai X, Gakidou E, Lopez AD. Evolution of the global smoking epidemic over the past half century: strengthening the evidence base for policy action. Tob Control 2022;31:129–137.

20. Mills KT, Stefanescu A, He J. The global epidemiology of hypertension. Nat Rev Nephrol 2020;16:223–237.

21. Forouzanfar MH, Liu P, Roth GA et al. Global Burden of Hypertension and Systolic Blood Pressure of at Least 110 to 115 mm Hg, 1990-2015. JAMA 2017;317:165–182.

22. Daviglus ML, Stamler J, Pirzada A et al. Favorable cardiovascular risk profile in young women and long-term risk of cardiovascular and all-cause mortality. JAMA 2004;292:1588–92.

23. Walli-Attaei M, Joseph P, Rosengren A et al. Variations between women and men in risk factors, treatments, cardiovascular disease incidence, and death in 27 high-income, middle-income, and low-income countries (PURE): a prospective cohort study. Lancet 2020;396:97–109.

24. Vaduganathan M, Mensah GA, Turco JV, Fuster V, Roth GA. The Global Burden of Cardiovascular Diseases and Risk: A Compass for Future Health. J Am Coll Cardiol 2022;80:2361–2371.

25. Schwalm JD, McCready T, Lopez-Jaramillo P et al. A community-based comprehensive intervention to reduce cardiovascular risk in hypertension (HOPE 4): a cluster-randomised controlled trial. Lancet 2019;394:1231–1242.

26. Sun Y, Mu J, Wang DW et al. A village doctor-led multifaceted intervention for blood pressure control in rural China: an open, cluster randomised trial. Lancet 2022;399:1964–1975.

27. Munoz D, Uzoije P, Reynolds C et al. Polypill for Cardiovascular Disease Prevention in an Underserved Population. N Engl J Med 2019;381:1114–1123.

28. Chow CK, Nguyen TN, Marschner S et al. Availability and affordability of medicines and cardiovascular outcomes in 21 high-income, middle-income and low-income countries. BMJ Glob Health 2020;5.

29. Kasim SS, Ibrahim N, Malek S et al. Validation of the general Framingham Risk Score (FRS), SCORE2, revised PCE and WHO CVD risk scores in an Asian population. Lancet Reg Health West Pac 2023;35:100742.

30. D’Agostino RB, Sr., Vasan RS, Pencina MJ et al. General cardiovascular risk profile for use in primary care: the Framingham Heart Study. Circulation 2008;117:743–53.

